# Adaptive sentinel testing in workplace for COVID-19 pandemic

**DOI:** 10.1101/2022.07.18.22277434

**Authors:** Yi Li, Mandy Chen, Joshy George, Edison T. Liu, R. Krishna Murthy Karuturi

## Abstract

Testing and isolation of infectious employees is one of the critical strategies to make the workplace safe during the pandemic for many organizations. Adaptive testing frequency reduces cost while keeping the pandemic under control at the workplace. However, most models aimed at estimating test frequencies were structured for municipalities or large organizations such as university campuses of highly mobile individuals. By contrast, the workplace exhibits distinct characteristics: employee positivity rate may be different from the local community because of rigorous protective measures at workplace, or self-selection of co-workers with common behavioral tendencies for adherence to pandemic mitigation guidelines. Moreover, dual exposure to COVID19 occurs at work and home that complicates transmission modelling, as does transmission tracing at the workplace. Hence, we developed *bi-modal SEIR model* and R-shiny tool that accounts for these differentiating factors to adaptively estimate the testing frequency for workplace. Our tool uses easily measurable parameters: community incidence rate, risks of acquiring infection from community and work-place, workforce size, and sensitivity of testing. Our model is best suited for moderate-sized organizations with low internal transmission rates, no-outward facing employees whose position demands frequent in-person interactions with the public, and low to medium population positivity rates. Simulations revealed that employee behavior in adherence to protective measures at work and in their community, and the onsite workforce size have large effects on testing frequency. Reducing workplace transmission rate through workplace mitigation protocols and higher sensitivity of the test deployed, though to a lesser extent. Furthermore, our simulations showed that sentinel testing leads to only marginal increase in the number of infections even for high community incidence rates, suggesting that this may be a cost-effective approach in future pandemics. We used our model to accurately guide testing regimen for three campuses of The Jackson Laboratory.

## 1 Introduction

The Jackson Laboratory (JAX) is a free-standing not-for-profit genetics research institute of ∼2,800 employees distributed in three major campuses in Maine, California, and Connecticut. JAX is unique in that it is a hybrid of a pure academic research institution, and a production unit providing mice and services to universities and pharmaceutical companies. In Mar’2020, the pandemic was gripping the world and lockdowns were being announced throughout the USA. Following the guidelines of the respective state governments, all three campuses of JAX were shut down. The immediate measures to reopen Jax campuses were reducing the onsite workforce size, requiring social distancing and mask-wearing as guided by CDC, and importantly, employee testing to identify and isolate infectious staff from the workplace and thereby minimizing consequential infections. When JAX initiated universal screening for COVID-19 infection to identify asymptomatic viral shedders, there was a vigorous debate as to how frequently should the workforce be tested. It was recognized that testing frequency has not only major cost implications and disruption of work processes, but also social consequences because of the reluctance of some members of the workforce to the discomfort of frequent testing. Using population modeling approaches, it had been recommended that the optimal testing frequency in a university setting should be every 3 days. Earlier in the pandemic, this was neither financially nor logistically feasible for a distributed workforce as for JAX. Moreover, the work environment in an institution that has production and service in addition to lab-based biomedical research is significantly different from a university comprising tens of thousands of young students who are highly mobile during the day and evening.

Therefore, we decided to develop an algorithmic approach to dynamically determine the optimal testing frequency that takes JAX’s employee behavior characteristics into account. As an experimental probe, the testing was initiated at weekly intervals to obtain baseline information on our three geographical sites, California, Maine, and Connecticut. Upon investigation, we found that population (community) positivity rates and employee sizes at each campus contributed to the very different incidences of positives at different campuses. In addition, the positivity rates among JAX employees were significantly lower than the population positivity rate in each of the respective state of the campus. We hypothesized that the demographics of JAX staff and an effective COVID19 education program at Jax led to behaviors that lowered exposure of JAX employees and their families to COVID19 resulting in lower positivity rates among JAX staff as compared to the local community rates. Another important characteristic of workplace spread of COVID19 is that the employees of an organization are exposed to COVID19 both at the workplace and at social setting when they return home. The exposure would depend on a variety of factors including masking and social distancing regulations at work, idiosyncratic employee behavior in personal social settings, and exposure potential of the employee’s family. Given that although these parameters change across campuses and over time, the trends were consistent for each worksite. Therefore, we wanted to have an adaptive testing program so that would take account of the consistent patterns we observed at each state site, and project the minimal testing frequency for optimal cost and health effects for the workforce.

Compartmental models [1-2,15], especially the variants of SEIR models [3-8,15], have been shown to be effective at estimating trends of epidemics. For dynamic populations, the floating population models or multi-group models [9-12] have been proposed in the literature. They are effective at capturing trends of an epidemic due to within-group spread as well as exposure to a few distinct homogeneous groups. The goal typically is to estimate trends in each homogeneous group and guide policies such as travel and closure. Though the multi-group compartmental models have been shown to be effective at estimating trends of the epidemic when multiple groups interact such as incorporating floating population, they are complex and do not control for the average rate of transmission of infection from an infectious employee nor account for specific characteristics of a workplace with limited populations (between 500-5000) and employees who are not outward-facing (i.e. whose job is direct contact with the general public). Therefore, workplace modeling is different from multi-group modeling in several ways including the goal, and the population, population flow, the parameters to be measured, and interventions to be deployed. The goal of the workplace model is to estimate the minimal testing frequency required to achieve stringent control of the number of infections at the workplace to be below a pre-chosen threshold, rather than just flattening the curve. Unlike multi-group models, there is no *j*^*th*^ cluster in the workplace model. Staff come to work from home, interact at the workplace and return to their homegroup; thus, they constitute a fixed sample of a group that acts as carriers of external infections to the workplace and carriers of workplace infections back to their homegroup. The workplace model needs to control a few well defined and controllable factors such as interaction among employees (e.g., masking, population size at work, social distancing), contact tracing, testing, and quarantining. Hence, a workplace model must account for exposure in the home group as well as at work while accounting for the workplace measures and community (aka population) parameters. Separate workplace modeling can help quantify the impact of workplace measures (e.g., testing, mask, social distancing, etc) on the transmission of infection and should be used in the modeling along with the population (home group) parameters such as community positivity rate and exposure rates. The dynamic nature of these population parameters can easily be accounted for through public sources and testing frequency estimates be adjusted. Assessing workforce COVID rates also helps adjust the impact of population positivity rates since employee exposure in outside social settings will be strongly affected by adherence to regulations governing the homegroup and to personal risk tolerances.

To accomplish this, we developed a *bi-modal SEIR (bSEIR) model* to model COVID19 incidence & transmission at workplace and developed an R-shiny based tool to estimate the optimal testing frequency of employees to control the number of active cases at workplace. The model is based on the number of onsite employees, population positivity rate in the respective county, the quantified relative risk of infection assuming exposure of employees to COVID19 outside the workplace (using a Hazard ratio or HR), empirical internal transmission rate through contact tracing efforts in the work-place, and sensitivity of testing. Given the recent waves of vaccinations and their varying vaccine efficacy rates, our model was adjusted to account for it. The tool allows modeling testing frequency for sentinel or cohort testing. This is a stratagem where a fixed proportion of workers would be tested at a particular time interval until the entire cohort has been tested. In our bSEIR modeling, we assume that the population risk of all employees be homogeneous as most onsite employees live typically within commutable distance from the workplace. We also assume that the interaction among the workers is limited only to the workplace i.e., infectious employees may infect their colleagues only at workplace even though some workers will have social interactions outside the workplace.

Our *bi-modal SEIR* (bSEIR) model allows simple modeling of dual exposure (within the workplace and at home) and testing frequency by accounting for easily measurable community and workplace parameters of epidemic incidence and transmission. The simulations show the importance of employee education and support to minimize their exposure outside the workplace in reducing the testing frequency required. Furthermore, sentinel testing, a logistically feasible testing paradigm, was evaluated and shown that it only marginally (2-6%) increases the total number of infections over a 3-month period. In addition, our simulations show that a moderate vaccination rate of 40-60% among employees coupled with reduced population positivity rates would have a multifold reduction in testing frequency requirements. Employee testing may not be required at very low population positivity rates if employees continued to exercise caution in social settings. We used our tool to generate weekly reports to forecast the expected numbers of infections and to recommend testing frequency for each campus of The Jackson Laboratory. Overall, our model helped our organization to minimize the cost of testing while controlling the epidemic at our workplace.

## 2 Methods

### 2.1 SEIR model for a closed community

SEIR (Susceptible, Exposed, Infectious and Removed) model [3-8] (Figure 1, top panel) is best suited to model pandemics in a closed community. As per an SEIR model, the community members belong to one of the four compartments: S*usceptible* (S) compartment where a subset of the community is not immune and not exposed to the virus in the current cycle, *Exposed* (E) compartment where another subset of members are infected with the virus but are not infectious yet, *Infectious* (I) compartment where infected members are infectious, *Recovered/Removed* (R) compartment holds the members who are infected and recovered from the infection (and become immune to the virus in this article) or succumbed to the infection. Assume N is the total number of people in the community, S, E, I, R are the number of people in the four compartments respectively at time t i.e., S+E+I+R = N. β, α, γ are the conversion rates from compartments S, E, and I to E, I, and R respectively. µ and Ʌ are the natural death and birth rates. The dynamics of the fraction of people in the four compartments are specified in the following system of differential equations and can be solved in a closed form under certain conditions [15].

**Figure 1:**
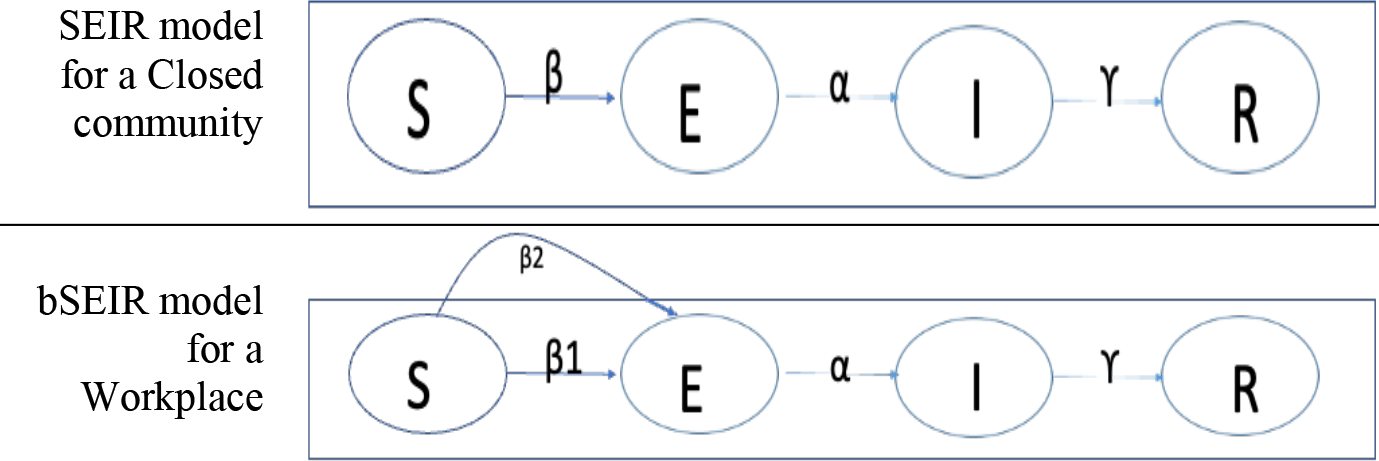
Modeling the pandemic: (top) SEIR model for the closed community, and (bottom) bi-modal SEIR model for the workplace (open system) - β1 and β2 are conversion rates from S to E due to exposure work and at home group respectively.

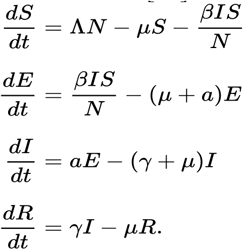

The conversion rates, β, α, γ, are determined by the model parameters (Tables 1 and 2 along with the Equation 1). For example, β, the conversion rate from S to E compartment, is determined by the average number of members an infectious member can infect during his/her infectious period within the community; α, the conversion rate from E to I compartment is determined by the average length of the latent period (LP) of the virus; γ, the conversion rate from I to R compartment is determined by the average number of days from being infectious to symptom manifestation (Pre) or when a person is tested positive and hence is isolated.

**Table 1:** Parameters characterizing the spread of the pandemic

| Parameter | Description | Default value |
| --- | --- | --- |
| p1 | the proportion of asymptomatic cases in SARS-CoV-2 cases | 0.5 |
| LP | SARS-CoV-2 virus latent period (days) | 3 |
| Pre | average no. of days being infectious before symptoms onset | 4 |
| Total | average no. of days an infectious person remains infectious | 14 |
| C | no of employees an infectious employee can infect in a workforce | 1 |

**Table 2:** Parameters in the *bSEIR* model. Choice or calculation of their values is addressed in Methods

| Parameter | Description | default value | comments |
| --- | --- | --- | --- |
| C | no of employees, an infectious employee can infect in a workforce | 1 | depends on the protective measures in the organization |
| Posrate<br>(Positivity rate) | no of positive cases per 1000 tested in the local community | daily positivity rate in the corresponding state or county |  |
| HR<br>(Hazard Ratio) | probability of exposure to the infections in the local community outside the workplace | 0.2 | empirically estimated using test data. (see methods) |
| I | no of infectious employees that are tolerated at a workforce | 1 | To be adjusted based on the organization characteristics (e.g. # of employees, nature of work, testing and social distancing measures) and population parameters, and feasibility |
| N | no of employees on-site | workplace specific |  |
| unit | time unit | week | Either week or day |
| contact-TracingEff | proportion of close contacts of a positive case that can be traced | 0 | We set it to 0 for a conservative estimate and due to low internal transmission rate |
| sens | test sensitivity | 0.95 | This should be adjusted based on the testing kits used. |
| spe | test specificity | 1 | This should be adjusted based on the testing kits used. |

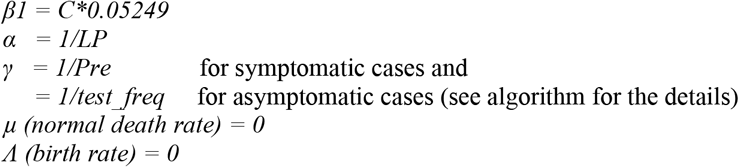

**Equation 1:** Computing SEIR conversion parameters

### 2.2 bi-modal SEIR (*bSEIR*) to model dual exposure open group

The SEIR model is applicable only to a closed community where a certain number of infected members are present at the beginning of a simulation, and SEIR models the trajectory of number of infections within this closed community. However, members (employees) of a typical workplace are exposed to COVID19 both within and outside the workplace on daily basis. Therefore, we extended the SEIR model to this setting (bSEIR model in Figure 1, the bottom panel) where susceptible employees are exposed both within (with the rate of β1) and outside (with the rate of β2) the workplace every day. The internal exposure rate, β1, depends on a workplace’s extant of pandemic and the protective measures adopted. The external exposure rate, β2, is determined by the probability of exposure to the community infections (hazard ratio or HR) and the local community or population positivity rate (P). The HR is estimated based on the empirical test data from the workplace, described in the following subsection. The *bSEIR* model accounts for the infections of employees from their home community as daily incoming waves of infections, followed by conversions from E to I and I to R as in an open community model. The *bSEIR* model is aimed at controlling the average number of employees in I, typically to be under 1 in our case, in small to moderate-sized organizations

We account for both symptomatic and asymptomatic types of COVID19 positive cases present in a workforce with a pre-specified proportion. We assume that asymptomatic positive cases are isolated upon being tested positive; while symptomatic cases are isolated upon symptom onset or tested positive before the symptoms manifest, whichever is earlier. The employees traced to be in close contact with COVID19 positive employees will be quarantined upon the identification of both types of cases. We also assume that the test can only detect infectious cases (with a specified sensitivity and specificity) but not all infected cases, and the infectiousness is uniformly distributed over the infectious period for both types of COVID19 positive cases. The assumptions are summarized in Table 3.

**Table 3:** Assumptions of Bi-modal SEIR model.

| Assumption | Impact | Range of parameters |
| --- | --- | --- |
| COVID19 tests are positive for infectious staff only. | Results in false negatives, will miss those in E, but not infectious. | Test sensitivity= 0.5 to 0.95. Our default value is 0.95 if PCR testing is used |
| Asymptomatic staff are identified by positive test results and are quarantined immediately | There is a delay between being tested and being quarantined. They could spread during this period. | Assuming there is no delay between a positive test result and quarantining. |
| symptomatic employees are identified either by symptoms or testing | no of days being infectious before symptoms onset varies among employees. Only the average period is accounted for in the model. | Mean number of days: 4<br>User can change this parameter from 3 to 7 |
| Fixed proportion of asymptomatic and symptomatic positive cases | Affects how people in I compartment move to R compartment | Default value: 0.5<br>User can change this parameter from 0 to 1 |
| Employees catch the virus 1) exposure to local population, 2) by colleagues at workplace | Resulted in bSEIR model. It doesn't account for variation among employees. Only averages are accounted for. | Three major parameters of bSEIR model are :1) pos-rate, 2) HR, and 3) C. pos-rate is projected for a future week based on historical data and HR is estimated using empirical data. |
| tests negative while being healthy or while incubating the virus (infected, not infectious yet) | It provides a conservative estimate of testing frequency. | The incubation parameter can be adjusted by the user. The default value is 3 |
| Infectiousness is uniformly distributed over the infectious period, and assumed to be the same for symptomatic and asymptomatic cases | This leads to higher-than-required testing frequency as infectiousness tappers off over the course of infection. | This can be handled by the user settable parameter *total*. The default value is 14 |
| Infection independence. Any two infectious employees infect a non-overlapping set of employees at the workplace | If this assumption is violated, the model estimates a higher number of infections than actual, leading to higher-than-required testing frequency | The internal transmission rate can be adjusted based on the tracking data at the workplace. |
| Low internal transmission rate. Assumes good protective measures: social distancing and masking are effectively imposed. | If the internal transmission rate goes higher, the independent infection assumptions will be violated and lead to a higher-than-required testing frequency. | The parameter C can be adjusted to account for the deviations from this assumption. |
| Employees are either fully onsite or fully off-site. All partial onsite employees are | Leads to higher-than-required testing frequency. | An organization can deal with it either by adjusting the effective number of employees by weighing |
| accounted as fully on-site for the modeling purposes and the modeling is performed only for on-site employees. |  | each employee proportional to their onsite time. |
| Infected employees won't be reinfected within the period monitored | Leads to lower-than-required testing frequency. | Users can keep the number of employees at the same level as the number of on-site employees at any time thus getting a conservative estimate of testing frequency. |
| The workforce is not outward facing | The outward-facing workforce may have a higher chance of being infected e.g., sales staff in grocery stores and healthcare staff. It can lead to higher HR and violating the independence assumptions. | The user can set HR to an appropriate level as data emerges with a good contact tracing. Our model can estimate HR based on observed data on infections and internal transmission rates. |

**Table 3:** Predictions available from the tool and the features of the tool

| Major predictions or features of the tool | Description | Main Parameters to set |
| --- | --- | --- |
| Predicting testing frequency | Weekly testing proportion | C, posRate, HR, N |
| Estimating spread of pandemic – given testing frequency. | Plot of number of infectious employees over a period | C, posRate, HR, N |
| Change alerts monitoring. | Whether the number of infectious people in recent tests exceed the upper bound; if yes, estimate new testing frequency to get I under control | If yes, set New Obs: no of tests to a positive number. if no, set it to 0. |

We implemented our *bSEIR* model using the deterministic implementation of the SEIR model in [2]. A user set parameter is available to control the number of infectious employees that can be tolerated throughout the staggered testing period in our online tool. The default values of the parameters of the progression of infection and occurrence of symptoms are obtained from [1]. The tool can recommend the minimal daily test proportion that satisfies the requirement of this parameter. The parameters along with their explanation and default values are given in Table 2. The assumptions of bSEIR model and their impact is explained in Table 3.

#### *bSEIR* Algorithm

Let us assume the following:

*SEIR(·)*: is an R function in the R package *EpiDynamics* [13] to estimate the fraction of employees in compartments S, E, I, and R at next day given the conversion rates *prop*: proportion of employees that are tested every day

*γ*_*1*_ (conversion rate from I to R for asymptomatic cases) = *prop*sens + (1- prop*sens)/total*

*γ*_*2*_ (conversion rate from I to R for symptomatic cases) = *prop*sens + (1-prop*sens)/Pre D*: number of days to estimate the fractions

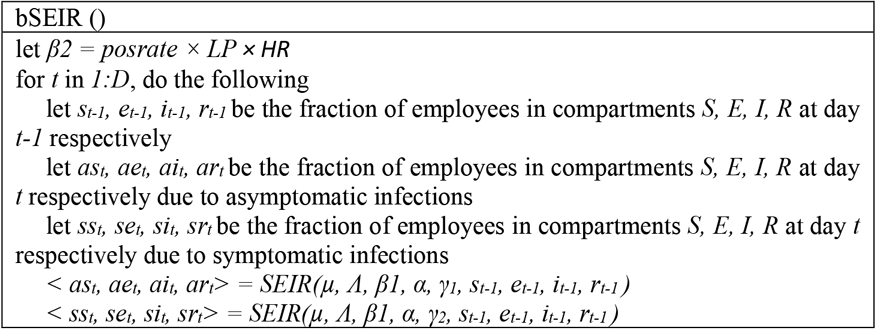

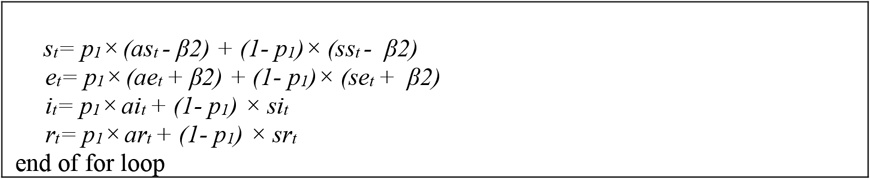

**Equation 2:** Algorithm for bSEIR Model

#### 2.2.1 Incorporating vaccination rate and efficacy in the bSEIR model

The vaccine rate (*Vr*) is defined as a fraction of employees fully vaccinated i.e., *Vr* = *(#of employees fully vaccinated)/(#of employees in the organization)*. If *Vri* is the proportion of employees received vaccine *i* (e.g., Moderna, Pfizer, J&J, etc) whose efficacy is *Vei*, then the overall efficacy of vaccination is estimated as weighted efficacy of all vaccine choices received by the workforce, as given below:

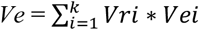

We can incorporate vaccination information in two different ways, by

1. Estimating the effective number of employees in S compartment i.e., *Se = S*(1-Ve)*. The underlying assumption is that *S*Ve* employees neither be infected nor be infectious, the assumption supported by recent studies [14]. or
2. Adjusting employee exposure to account for vaccinated employees i.e. *HRe = HR*(1- Ve)*^*½*^. The underlying assumption is that the vaccinated employees’ exposure to COVID will not lead to being infectious.

Of these two choices, we use (1) for its simplicity and ease of estimation.

#### 2.2.2 Hazard ratio (HR) estimation

##### Method 1

The hazard ratio can be estimated using empirical employee testing data. The (backward) 14-day moving average positivity rate for day *t* (*L*_*t*_) was calculated based on the testing data from the Jackson Laboratory. The number of new tests and the number of new positive tests each day for each state were downloaded from https://covidtracking.com/data/download, and the (backward) state-specific14-day moving average positivity rate for day *t* (*P*_t_) was calculated. The hazard ratio *H*_*t*_ at day *t* is computed as *L*_*t*_*/P*_*t*._ Since the number of employees at each campus is small, *L*_*t*_, hence, *H*_*t*_, are zero when *P*_*t*_ is below 5% (Figure 3). We calculated the average over non-zero *H*_*t*_ for a campus and used it as the probability of exposure to the local population risk. As shown in Figure 3, *H*_*t*_ estimates are not significantly different among campuses

##### Method 2

In another method, we estimated the number of infected persons in a week based on our simulation model and searched the best HR which minimized the mean squared difference between the observed and predicted number of infected persons in a week. To increase the number of data points to estimate HR, we assumed that the three campuses had the same HR and pooled the estimation data points from the three campuses together. This method is useful when the incidence rates are low.

#### 2.2.3 Community positivity rate projection

To estimate the testing frequency for a future week, the average daily population or local community positivity rate for a future week is needed besides other parameters. We assumed the daily positivity rate changed smoothly with time and estimated the daily population positivity rates for the next 7 days by polynomial curve (with a degree of 2 or 3) fitting of the observed daily population positivity rates during the past few months, followed by averaging on the 7 projections.

## 3 The Tool (https://rshiny.jax.org/connect/#/apps/156/access)

We developed an R-shiny based tool for interactive analysis to predict infected number of employees at a future time point, to check whether testing frequency to be increased, or to estimate testing frequency. We showed the functions of our tool in Table 4 and screenshots of the interfaces in Figure 2. The parameters associated with the workplace are placed on the left panel of the interface and the parameters associated with the spread of the pandemic are placed on the right panel of the interface for ease of operation.

**Table 4:** Detectability of COVID-19 positive cases

| Community Positivity rate | Min #of employees at power=0.7 | Min #of employees at power=0.85 |
| --- | --- | --- |
| 0.5% | 540 | 760 |
| 1% | 269 | 379 |
| 2% | 134 | 188 |
| 5% | 53 | 74 |
| 10% | 26 | 36 |

**Figure 2:**
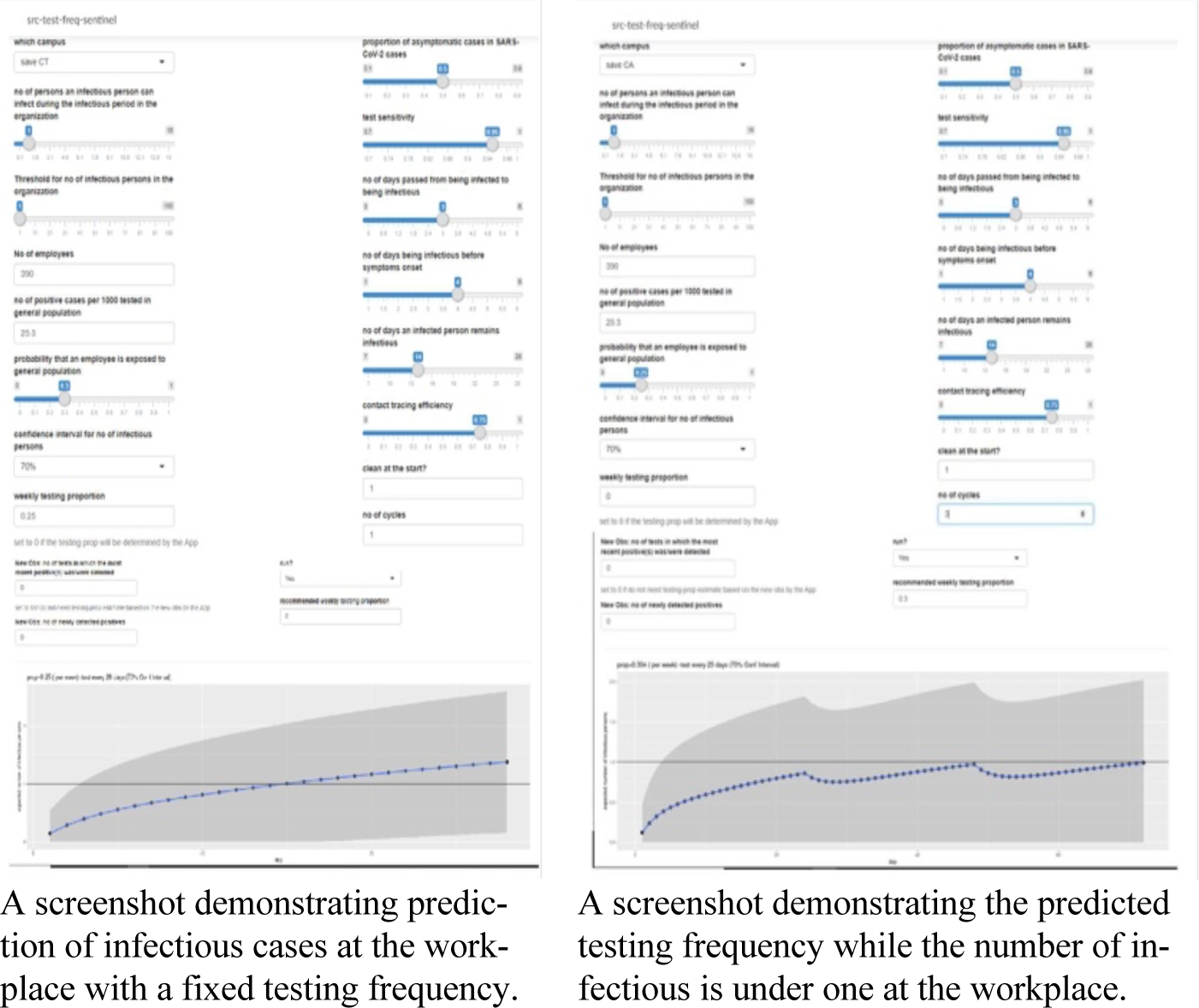
bSEIR R-Shiny tool - The R-shiny interface of the bSEIR tool. Screenshots of bSEIR tool for pandemic spread at workplace (left) and estimating testing frequency (right).

## 4 Results

### 4.1 COVID19 positive cases are detectable even in organizations as small as 300 employees but depends on community positivity rate

We conducted a power analysis of our model to identify the occurrence of a single infection among a workforce. Figure 3 shows the plots of a minimum size of the organization required for different population positivity rates, for different powers of detection. The same data is shown in Table 4 for the power of detection at 70% and 85%. At a population positivity rate of as low as 1%, we can detect the occurrence of one infection in an organization of size as small as 300 at a power of 80%. This is critical for the testing program to be effective at workplace.

**Figure 3:**
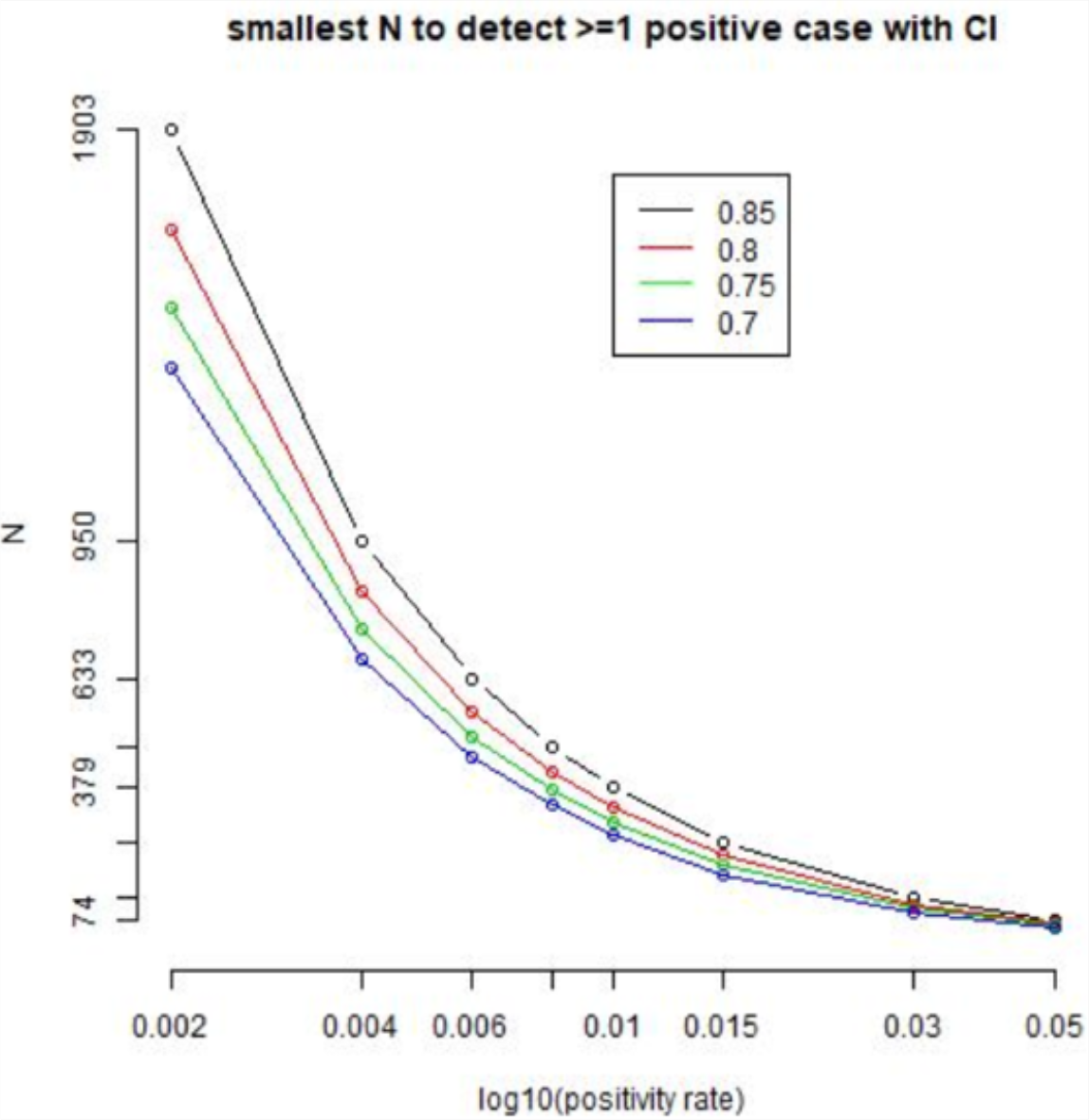
Organization size (N) required to detect at least one positive case depends on population positivity rate. Four colors stand for detection power of 0.85, 0.8, 0.75, 0.7 respectively.

### 4.2 Size of the workforce and employee behavior coupled with community positivity rate have a major impact on the frequency of testing

We conducted simulations to assess the influence of population and workplace parameters on the testing frequency required to control the number of infectious staff (I) to be <=1 at the workplace. The simulations used *contact tracing efficiency = 0, clean start, monitor from Day1, test sensitivity=0*.*95, without a gap between sentinel testing cycles*. The results (figure 4) show that the frequency of testing mostly depends on *hazard ratio (HR), size of the workforce* and *daily population positivity rate*. Small to moderate internal infection rates have significant effect on testing frequency for lower population positivity rates or smaller hazard ratios. Large organizations (no of employees ∼5000) need to keep hazard ratio small even for small population positivity rates to keep the testing frequency lower to avoid outbreak at the organization. Frequency of testing is significantly affected by the size of the organization and hazard ratio, while internal transmission rate has a significant impact on testing frequency only when low testing frequency is possible i.e., for low HR and low community positivity rate. The sensitivity of testing also has marginal effect on testing frequency (data not shown here).

**Figure 4:**
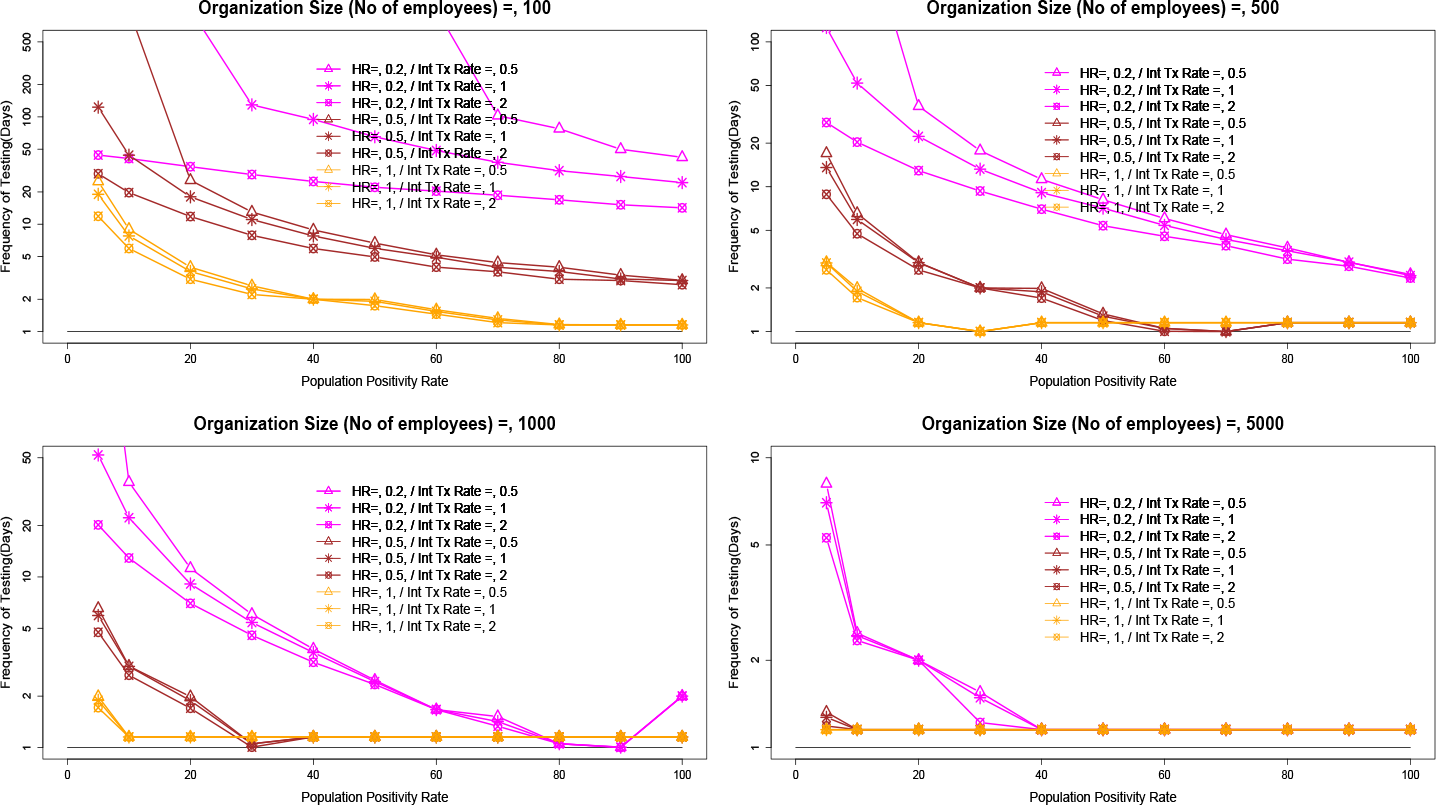
Frequency of testing to control the number of infectious employees to be <=1 (test sensitivity = 0.95, clear start of the cohort, no tracking and monitor the cases from day 1, no gap in testing). The frequency of testing mostly depends on the hazard ratio and size of the organization. Small to moderate internal infection rates have a significant effect on testing frequency for lower population positivity rates (per 1000) and smaller hazard ratios. Moderate-sized organizations (EE#=5000) need to keep hazard ratio small even for small population positivity rates to control outbreaks at the organization.

### 4.3 Sentinel testing leads to marginally higher infections than cohort testing at higher HR (Contact tracing efficiency = 0, clean start, monitor from Day1)

Sentinel testing, a schedule of testing all employees over an extended period, is typically adopted due to its feasibility of logistics and resource availability for testing. For example, for a cohort of 400, the population can be tested in blocks of 100 per week until all 400 individuals have been tested at least once in the 4-week interval. However, we may expect it to lead to higher number of infections compared to testing all employees at one go i.e., cohort testing. Our simulations (figure 5) show that, over a 90-day period, sentinel testing leads to only a marginally higher (2-6% more i.e., fold change of 1.02-1.06) infections than cohort testing. The impact will be lower if the hazard ratios and population positivity rates are low. Sentinel testing can be further divided into two categories, one without gaps between testing cycles and the other allowing variable proportion of testing with fixed gaps. The former results in a lower number of infections but much higher relative cost, while the later leads to higher average proportion of testing but lower number of total employees infected (data not shown).

**Figure 5:**
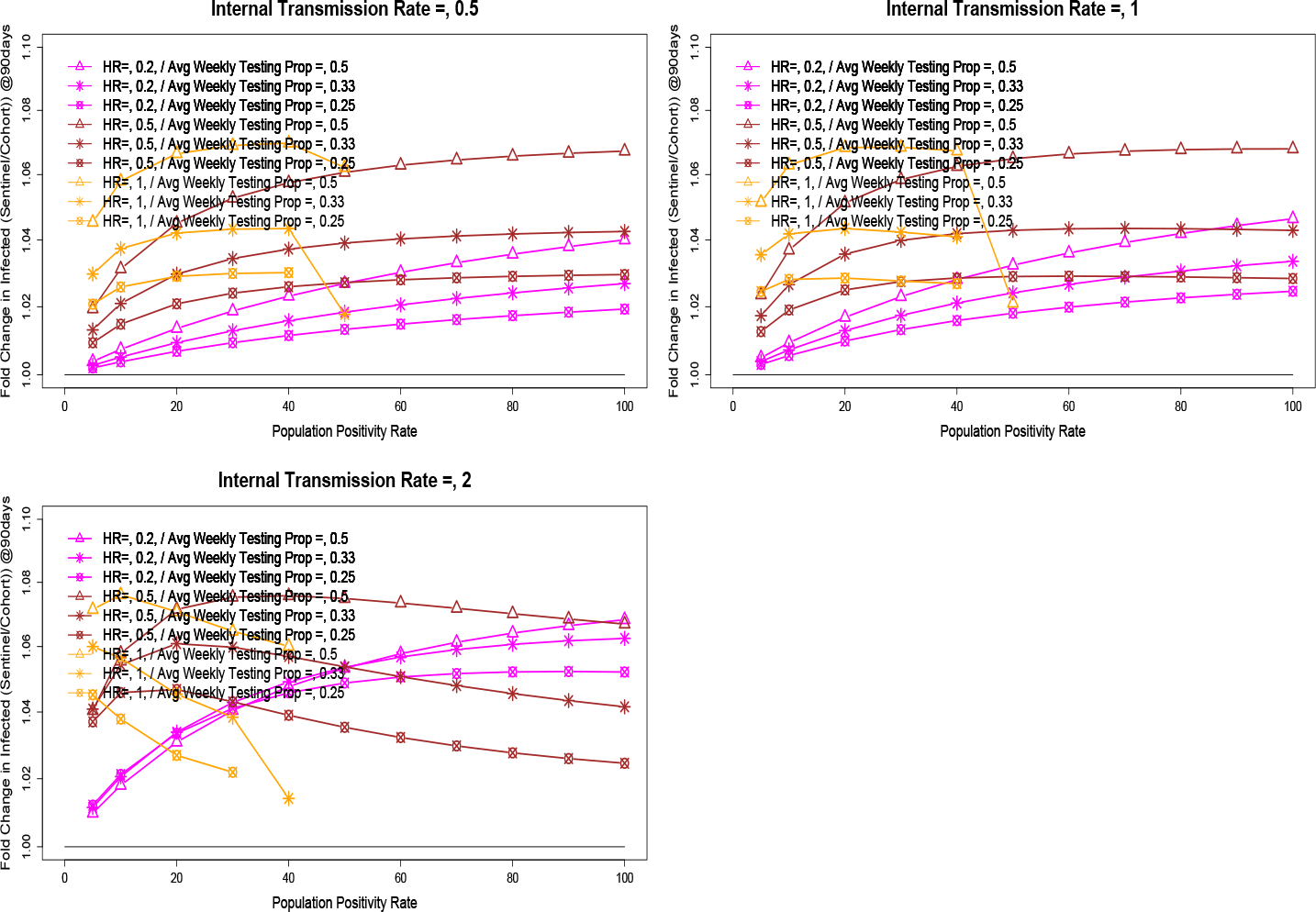
Ratio of Frequency of testing for sentinel testing vs cohort testing for a fixed average weekly proportion of testing of 0.25, 0.33 and 0.5. The analysis is based on test sensitivity = 0.95 and Organizational size=500.

### 4.4 Impact of Vaccination Rate on Testing frequency

We conducted simulations to understand the impact of efficacy rates of vaccines against COVID19 infections on the testing frequency. We used both models discussed in Methods section to account for vaccination rates in our bSEIR model i.e., adjusting for HR vs adjusting for effective population. Though both models give rise to similar testing frequency, adjusting for HR (y-axis) leads to higher frequency of testing (sup figure *Vac*) for a subset of scenarios.

Even a modest vaccination rate has a significant effect on the testing frequency, especially for medium organizations of size below 1000 employees. The impact is extremely high for low HR and medium population positivity rates, as highlighted by the cyan-colored lines in Figure 6.

**Figure 6:**
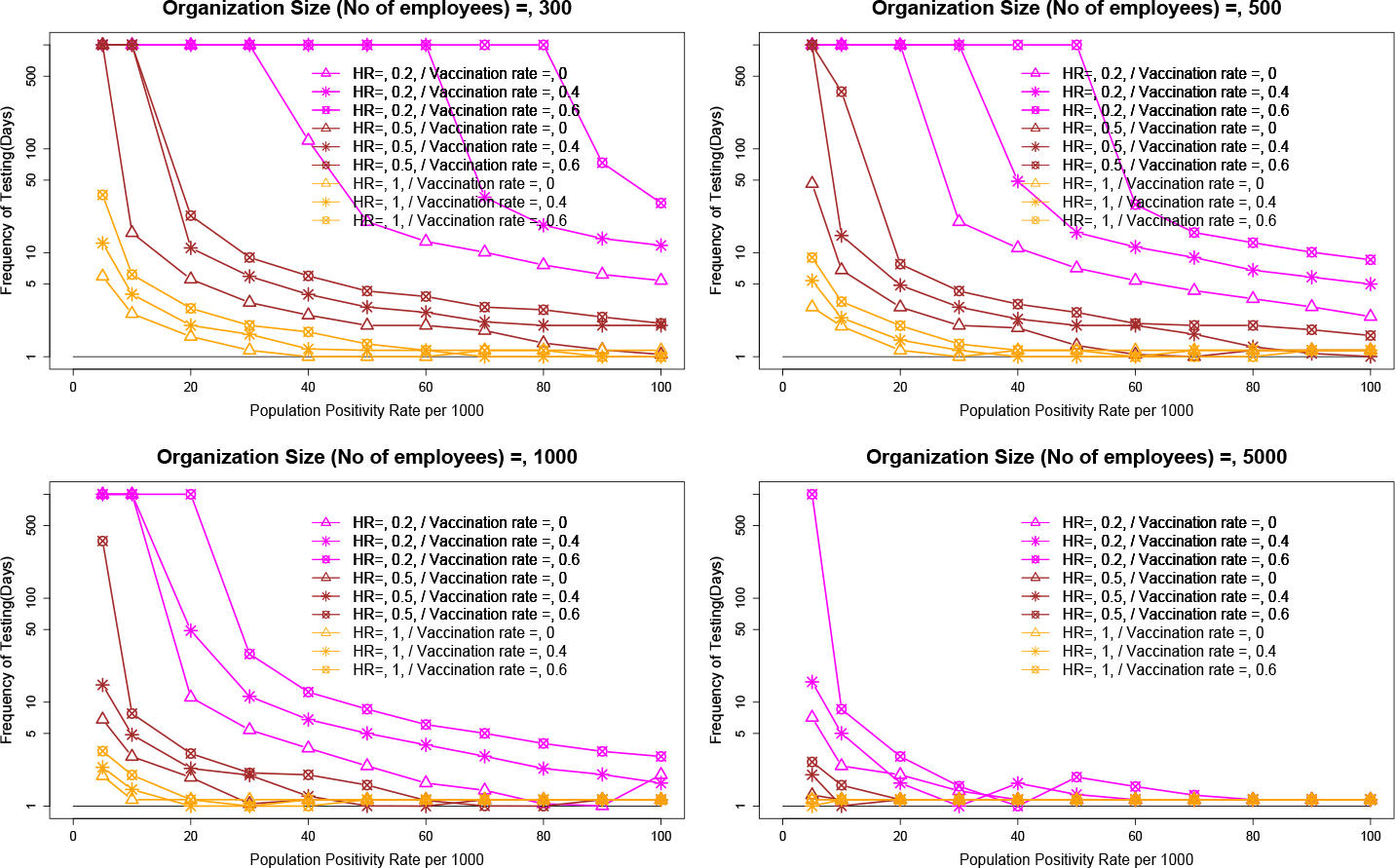
A modest vaccination rate of 40-60% has a huge impact on testing frequency, especially at low population positivity rates or a low hazard ratio of 0.2. Therefore, population vaccination, employee vaccination, and continued education of employees are important factors to keep the testing frequency low for the economics of organizations and the health of the employees.

### 4.5 Adaptive Sentinel Testing at JAX - An application of our tool

To apply bSEIR model for Jax’s adaptive sentinel testing policy, we projected community positivity rates and hazard ratios for all campuses of Jax.

Figure 7 shows that our projections of community positivity rate are within 20% error for most of the time, for all campuses. However, the relative error is higher for lower community positivity rates, mostly overestimating the community positivity rate which results in a higher-than-required testing frequency only for mid to higher HR at low community positivity rates.

**Figure 7:**
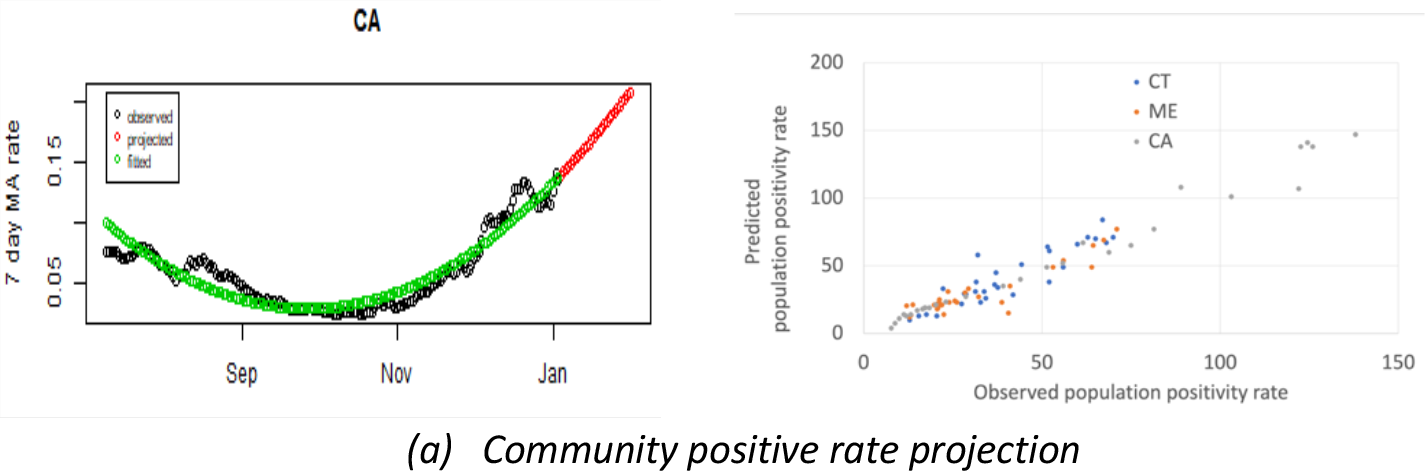
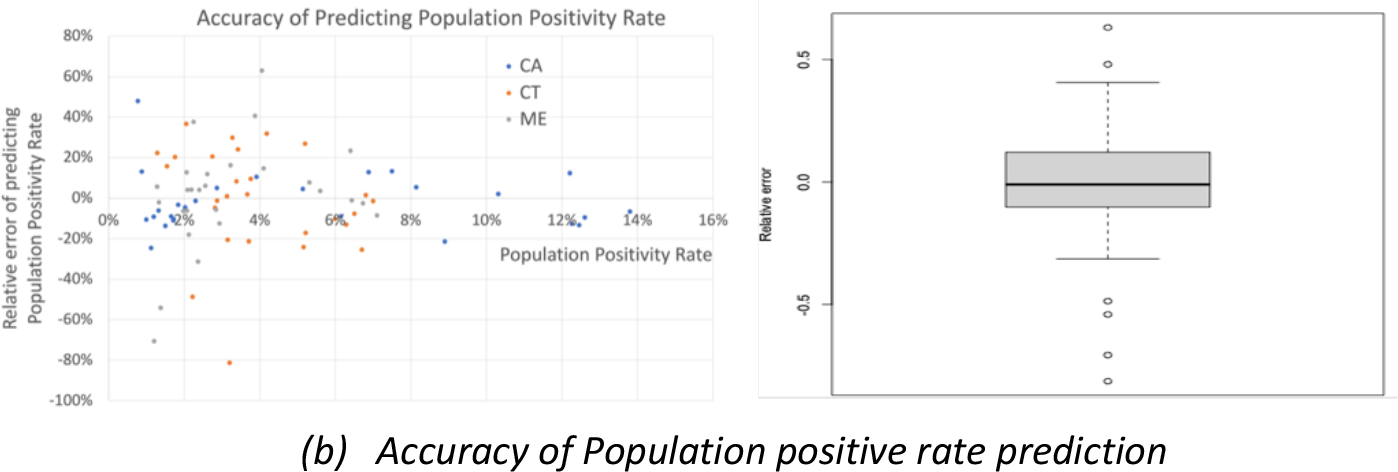
Projecting community positive rate using a polynomial model fitting, and its accuracy across Jax campuses.

Typical HR turned out to be 0.25 for all Jax campuses. However, the HR was expected to spike during the holiday season of Thanksgiving and Christmas in 2020. Hence, we set HR for normal weeks (=0.25) and holiday weeks (=0.35).

The scatter plot in figure 8 shows a very strong correlation (=0.68) between our prediction of the number of cases to the observed number of cases.

**Figure 8:**
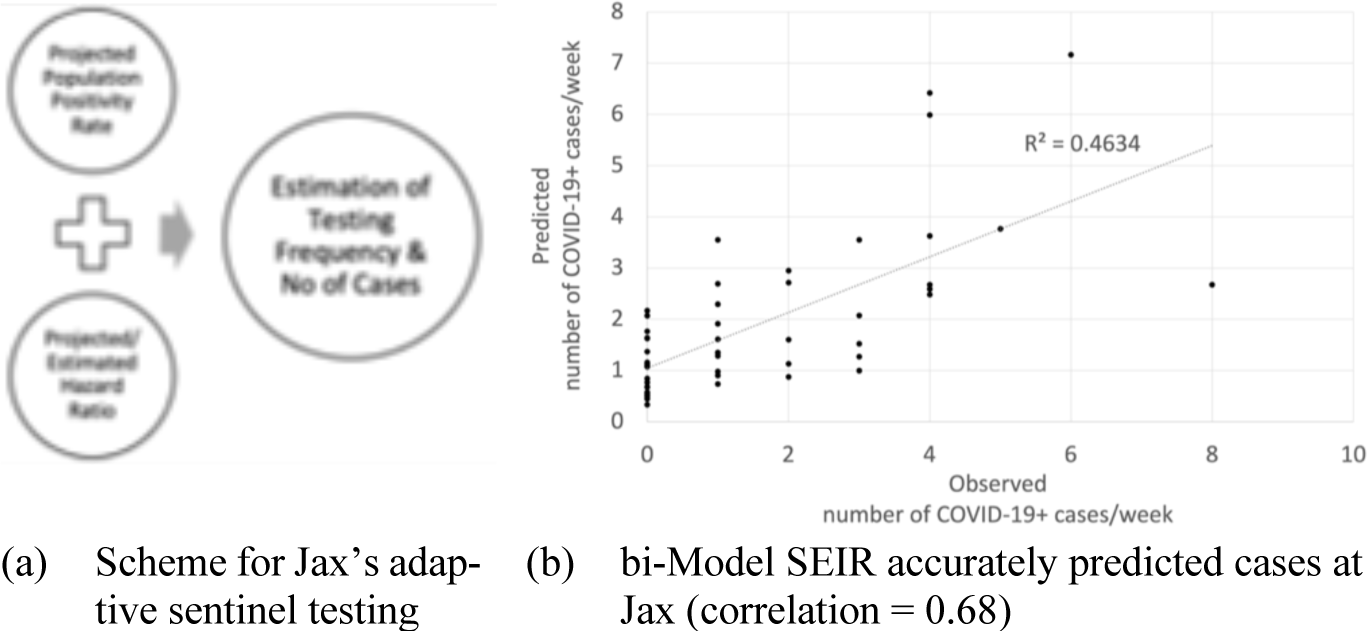
Performance of Bi-Modal SEIR model.

## 5 Discussion

Adaptive testing frequency is crucial to balance between economics of testing and to keep the pandemic under control at the workplace. We developed a *bi-modal SEIR* (*bSEIR*) model to account for changing parameters of the pandemic in the community and the workplace where the workforce is not outwardly facing.

Our *bi-modal SEIR* allows simple modeling of dual exposure, outbreak, and testing frequency by accounting for easily measurable community, and workplace parameters of epidemic incidence and transmission. The simulations show the importance of employee education and support to minimize their exposure to community infections in reducing the testing frequency required. Even a modest vaccination rate of 40-60% with vaccines of the efficacy of more than 90% would greatly reduce testing frequency by 2-4-fold. It is highly pronounced for low HR and medium to high community positivity rates. Furthermore, sentinel testing, a logistically feasible testing paradigm, was evaluated and shown that it only marginally (2-6%) increases the total number of infections over a 3-month period. Thus, sentinel testing is a viable cost-effective approach to manage testing in the workplace. We used our tool to generate weekly reports of forecast of population positivity rates, expected number of infections, and recommended testing frequency for The Jackson Laboratory. Overall, our model helped our organization to minimize the cost of testing while controlling the epidemic.

An apparent limitation of our model is that it does not explicitly assume the possibility of break-through infections which requires a fraction of employees in the *R* compartment to be moved to the *S* compartment. However, as we repeat the simulations with S to be total onsite workforce size and adjusting the vaccine efficacy rates to account for changing efficacy of vaccines with time, the break-through infections are indirectly accounted for which will be good enough to predict the short-term course of the pandemic at the workplace. Our formulation, to compute average vaccine efficacy, can be generalized not only for different types of vaccines, but also for different efficacies due to new variants and different time periods after the most recent vaccinations. Furthermore, our model can be applied for a higher internal transmission rate by adjusting the internal transmission rate that accounts for one employee being exposed to multiple infectious employees. Overall, the apparent limitations of the model can be dealt with appropriate adjustment or estimation of its parameters.

## Data Availability

All data produced and software developed in the present study are available upon reasonable request to the authors

## Notes

### Competing Interest Statement

The authors have declared no competing interest.

### Funding Statement

This study was funded by The Jackson Laboratory, USA

### Author Declarations

IRB of The Jackson Laboratory determined on November 16th, 2020, before any research procedures, that the study is Exempt human subjects research.

